# Experimental Device to Evaluate Aerosol Dispersion in Venues

**DOI:** 10.1101/2023.06.13.23291335

**Authors:** Isabell Schulz, Felix Hehnen, Kevin Harry Lausch, Sophia Mareike Geisler, Ümit Hasirci, Sophia Wolff, Tim Rese, Sebastian Schimek, Michael Lommel, Christian Oliver Paschereit, Stefan Moritz, Martin Kriegel, Ulrich Kertzscher

## Abstract

The COVID-19 pandemic has focused attention to the importance of understanding and mitigating the airborne transmission of pathogens in indoor environments. This study investigated the aerosol distribution in different indoor venues with varying ventilation concepts, including displacement, mixed, and natural ventilation. A measurement system was developed to investigate venue-specific aerosol distribution patterns using a sodium chloride solution as a tracer. To analyse the spatial dispersion of aerosols, Computational Fluid Dynamics (CFD) simulations were conducted in addition to experimental investigations. The investigations indicated the lowest aerosol load for the venue with displacement ventilation and the highest for the natural ventilated venue. The measurement system developed in this study provides a useful tool for assessing the effectiveness of ventilation measures in reducing airborne transmission of pathogens in indoor environments. It also proved its wide range of applications, as it can be used in various sized and shaped indoor environments, with or without an audience.

## INTRODUCTION

The COVID-19 pandemic caused a significant public interest in reducing the risk of airborne transmission of pathogens, stimulating the investigation of the emission and transport of respiratory aerosols indoors.^1^ The SARS-COV-2 virus can be transmitted via aerosols released by breathing, speaking, sneezing and coughing. The most common place for virus spreading are indoor environments^2^ and the respiratory route is the primary mode of virus transmission.^3^ The highest risk of an infection carry aerosols and droplets with a size of 2-10 μm, because these size droplets^4,5^ have negligible sink velocities and follow any airflows accurately.^6,7^ After evaporation they can linger in still air for up to 60 min.^8^

It is known that the implementation of appropriate ventilation measures can significantly reduce the risk of indoor infections.^9^ Displacement ventilation aims to keep the fresh and polluted air separated.^10^ A stream of fresh air from the floor is used to displace the waste air, which rises to the ceiling by buoyant force and is then removed from the room via air vents.^11,12^ This creates two levels of air, with cool fresh air in the occupied lower part of the room, and warm waste air in the unoccupied upper part of the room. Natural convection generates a cooling effect for the occupants, as the cool air rises and adsorbs the heat from them, which is then expelled from the room.^13^ In a mixing ventilation system fresh air is supplied to the room with a high initial mean velocity, creating a high turbulence intensity to promote good mixing of fresh and polluted air and an even distribution of temperature and pollutants throughout the venue.^12,14^ Natural ventilation is a ventilation method where natural forces, such as wind and thermal buoyancy force, drive outdoor air through purpose-built building envelope openings. ^15^ It is a ventilation method with very low investment and operating costs compared to mechanical ventilation systems. In addition, the effectiveness of natural ventilation on the infection risk potential and climatic conditions in the room is highly dependent on the outside temperature, the wind direction and the user.^16^

Developing methods to assess the risk of airborne pathogen transmission is essential to ensure that technologies used to control and improve air quality work effectively and efficiently.^1^ For individualized considerations and high levels of accuracy, experimental studies must be conducted to examine the efficacy of ventilation measures.^9^ Several strategies have been proposed for investigating aerosol distribution in indoor venues, each with their own advantages and limitations.

One method to directly detect SARS-CoV-2 in the air is active indoor air sampling followed by polymerase chain reaction (PCR) detection as recently shown by Dinoi et al.^17^ While this method attempts to capture the actual viral load under real-world conditions, it does not provide controlled boundary conditions.

Another method to evaluate the spread of aerosols is the use of tracer gases. A tracer gas is emitted into the venue and local concentrations are measured using infrared spectrometers.^9^ This measurement technique is convenient for both small and big venues with outside air exchange. However, the room needs to be completely free from the tracer gas of potential previous measurements to avoid contamination. Also, tracer gases cannot be filtered, making this method unsuitable for evaluating air purifiers that rely on filtration.^9^ Another method to evaluate the spread of aerosols is the use of tracer gases. A tracer gas is emitted into the venue and local concentrations are measured using infrared spectrometers.^9^ This measurement technique is convenient for both small and big venues with outside air exchange. However, the room needs to be completely free from the tracer gas of potential previous measurements to avoid contamination. Also, tracer gases cannot be filtered, making this method unsuitable for evaluating air purifiers that rely on filtration.^9^

A third method to observe how particles are transported in indoor environments is the surrogate particle method, where a particle generator is used for the emission of surrogate particles. Using optical particle counters or scanning mobility particle sizers at several positions allows the spatial and temporal measurement of particle number, concentration, and size distribution. For this, the room to be studied needs to be initially free of particles and all potential sources of particles, including humans. Consequently, this technique is not suitable for use in everyday environments.^9^

Lommel & Froese et al.^18^ developed a portable measurement system consisting of one aerosol generator and several aerosol absorbers. The aerosol generator continuously releases a 10% sodium chloride (NaCl) solution. The NaCl in the aerosol particles serves as a tracer, simulating the spread of an airborne virus. The aerosol captured by the absorber is dissolved in ultrapure water and the amount of NaCl is calculated via conductivity measurement. Thus, the system can measure the temporal and spatial dispersion of the aerosol. It can be used to examine the effects of protective measures like active or passive ventilation and face masks without underlying assumptions or constraints like clean room environments.

In this paper, the measurement system of Lommel et al.^18^ was modified. To investigate venue-specific aerosol distribution patterns, venues with different room characteristics and ventilation concepts were examined using the experimental measurement system and Computational Fluid Dynamics (CFD) analyses. Three venues with different ventilation concepts are in the scope of this paper, including displacement ventilation, mixing ventilation, and natural ventilation. The paper of Geisler & Lausch et al.^19^ used the data of this study to investigate the infection risk potential in the venues, asses the effect of different mitigation measures and develop recommendations. In this paper, the measurement system of Lommel & Froese et al.^18^ was modified. To investigate venue-specific aerosol distribution patterns, venues with different room characteristics and ventilation concepts were examined using the experimental measurement system and Computational Fluid Dynamics (CFD) analyses. Three venues with different ventilation concepts are in the scope of this paper, including displacement ventilation, mixing ventilation, and natural ventilation. The paper of Geisler & Lausch et al.^19^ used the data of this study to investigate the infection risk potential in the venues, asses the effect of different mitigation measures and develop recommendations.

## MATERIALS AND METHODES

The study of venue-specific aerosol distribution patterns was examined by experimental and numerical measurements. The experimental investigation was conducted with an aerosol measurement system which consists of an aerosol generator and seven portable absorbers.

### Aerosol generator

The aerosol generator used for this study is the Atomizer Aerosol Generator ATM 230 (Topas GmbH, Dresden, Germany). The aerosol generator has an external compressed air supply (FK65 pro, 9 bar max, Weldinger, Germany) and operates at 1.5 bar. The size distribution was measured by a phase doppler anemometry (PDA)^20^, and the data was fitted through a t-location-scale distribution, which is useful for modelling data distributions with heavier tails than the normal distribution. The emitted aerosols have a mean diameter of 2.4 μm with a standard deviation of 1.1 μm. The tracer aerosol consists of 10% NaCl in water. The sodium chloride within the emitted aerosol acts as a tracer particle. The aerosol is released with a controllable mass flow of 0.43 g min^-1^. The aerosol generator operates in a continuous emission mode. The tracer solution is stored in a separate 5 l container and is pumped in a circuit between the atomizer container (500 ml filling volume) of the aerosol generator and the storage container with the aid of a peristaltic pump (DC 12 V, Vikye, China). This ensures that the tracer solution has the same salt concentration throughout the dispersion and that the salt concentration of the tracer solution does not increase noticeably due to salt crystals which remain attached to the edge of the container. The tracer solution in the container is constantly heated to 37 °C.

### Absorbers

The absorbers inhale the ambient air and measure the amount of inhaled NaCl. The design of the absorbers is illustrated in Figure 1. A vacuum pump (DC 12V 12W V, VN-C3 Mini, Vikye, China) generates a defined continuous volume flow through an inhalation tube. The volume of the aspiration airflow is 10 l min^-1^. A fine filter (original coffee filter 1×6, Melitta, Germany) is attached to the end of the inhalation tube, surrounded by 150 ml of deionized water within a glass reservoir. The filter acts as an atomizer, breaking down the air stream into fine bubbles to dissolve the NaCl particles in the water. A peristaltic pump (DC 12 V, Vikye, China) ensures that the filter is constantly flushed, by pumping the water through a parallel circuit, in which a conductivity sensor (HI98192, Hanna Instruments, Germany) is located. The conductivity sensor has a resolution of 0.01 μS/cm and a measurement accuracy of ± 1%. The solved NaCl causes an increase of conductivity of the ultrapure water, enabling the determination of the NaCl amount.

**Figure 1:**
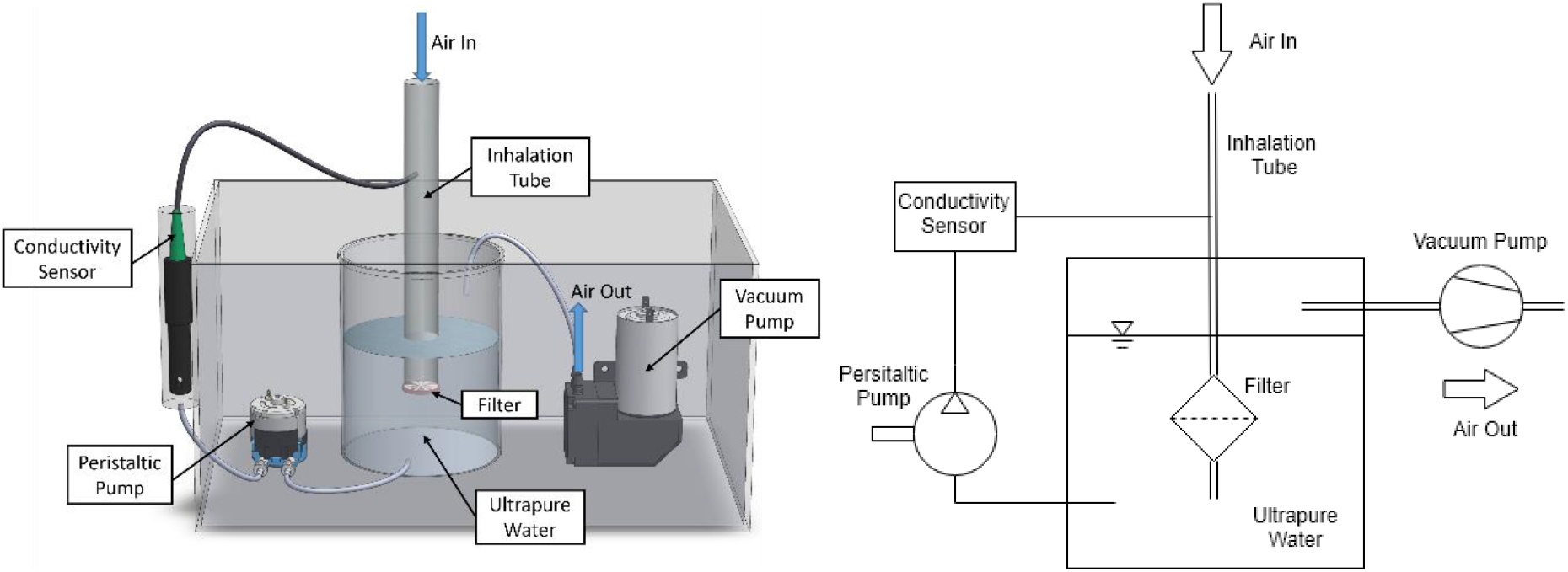
Illustration of an absorber: the aerosol is sucked through the inhalation tube into a closed reservoir filled with ultrapure water by a vacuum pump. The filter attached to the inhalation tube dissolves the NaCl particles in the ultrapure water. The ultrapure water is pumped through the sensor by a peristaltic pump.

### Measurement procedure

The measurements start with a lead-in time (I), which records current indoor air conditions in the absence of aerosol emission by measuring the increase in conductivity inside the absorbers. Thus, in the first minutes of the measurement, the background concentration of conductive material in the air is measured. This slight increase in conductivity caused by the room specific conductive aerosols or aerosols from previous measurements must be subtracted from the current measurement as an offset from the measured values during the post-processing. The lead-in time is also a good indicator of whether an environment has been sufficiently ventilated after previous measurements. Following the lead-in phase, the aerosol generator starts to release the tracer aerosols for a predetermined period of time. This phase is called the emission period (II). After the emission stops, the measurement is continued up to a predetermined time, called the lead-out time (III). During the entire measurement procedure (lead-in, emission, lead-out), which is displayed in Figure 2, the conductivity measurement of each individual absorber is recorded in real time on the measuring device. The conductivity sensor also continuously measures the temperature of the measuring fluid. The dependence of the conductivity on the temperature is corrected in the post-processing of the data. The room temperature and humidity are recorded accordingly.

**Figure 2:**
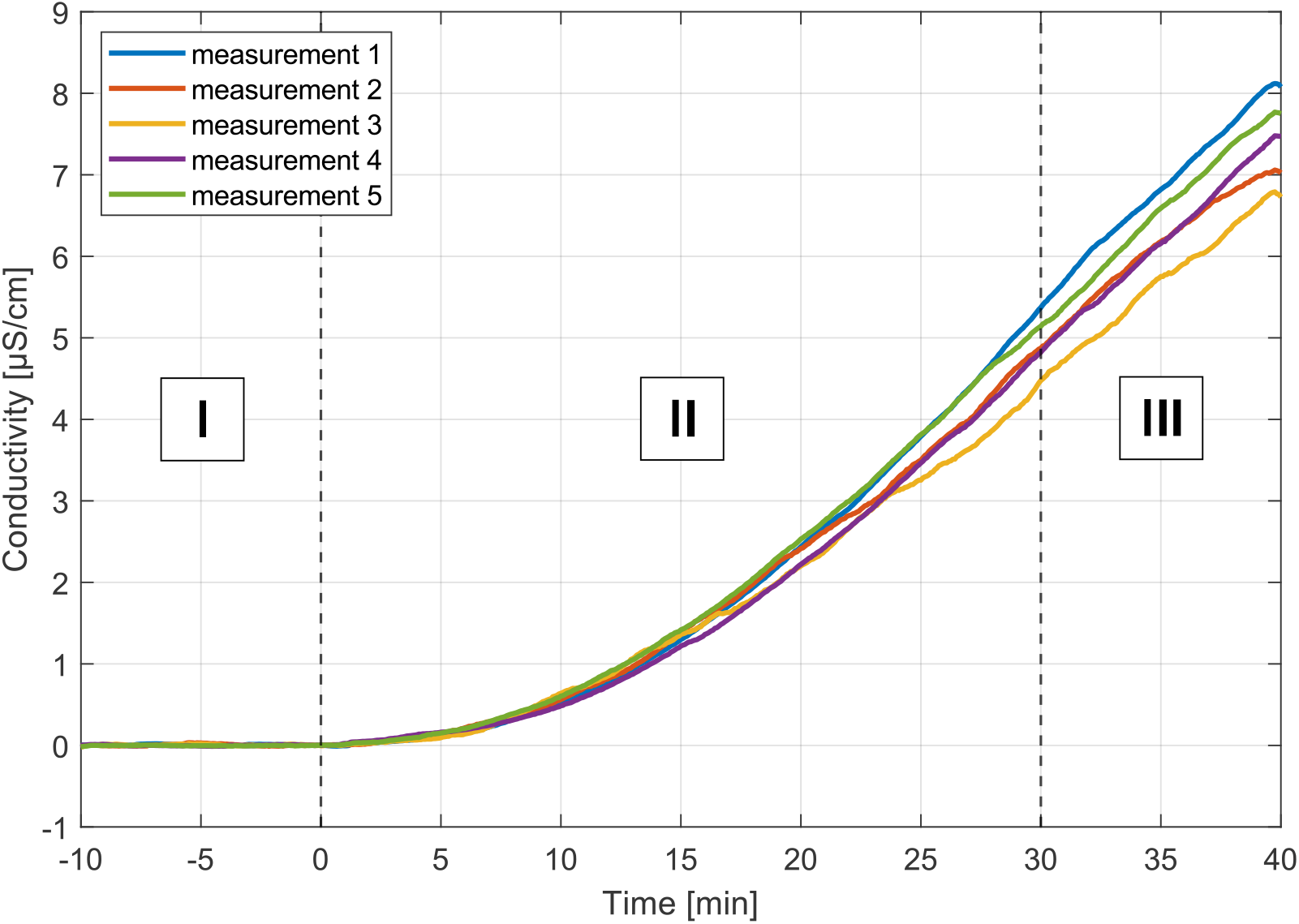
Increase in conductivity over time during a measurement procedure. The measurement procedure can be divided into three periods: lead-in (I), emission (II) and lead-out (III). Each measurement shows the mean value of the conductivity of all absorbers in the room. The lead-in time starts 10 min before the release of the aerosol. Then aerosol is released for 30 min and an increase in conductivity in the absorber can be observed. After 30 min the aerosol release stops, and the increase of the conductivity slows down.

The duration of the measurements at the individual venues varied, as the measurements were adapted to event-specific characteristics.

### Measurement setup

The aerosol generator and absorbers are positioned in different spatial arrangements to each other within the considered venue. The experimentally investigated seats were selected prior to the start of the CFD investigations and were chosen to the best of our knowledge to represent the aerosol distribution at the venue.

Venues can vary in terms of size and shape, ventilation system, and arrangement of the audience. For smaller venues (up to 100 seats) the measurements were conducted with heat sources to mimic the buoyancy effects generated by humans, which significantly influence the room airflow. For an appropriate simulation of the volumetric flow rate of a human thermal plume (75m^3^/h)^21^, heat sources were assembled consisting of a heat cable (80 W, Terra Exotica, Germany) and a bucket that is placed on the heating cable with the opening facing downwards. The heat output corresponds to the physiological human heat emission for a low activity rate (80W).^22,23^ The aerosol generator and the absorbers were placed on a heat source, whether measuring with or without audience.

### Reproducibility measurements of the system

To prove that the system can reproducibly measure aerosol levels in rooms, the same measurement was repeated five times in a sealed conference room. This room has a volume of 100 m^3^ and negligible air exchange. Two heaters constantly heat the room to 20 ± 1° C. Due to thermal convection, there is a constant mixing of the room air in the entire room. Four absorbers were positioned at 1.5 m around the aerosol generator. The reproducibility measurement followed the measurement procedure stated above. A lead-in time of 10 min was chosen to sufficiently measure the background concentration of the NaCl particles. The aerosol was released by the aerosol generator for 30 min to ensure a high signal-to-noise ratio. The lead-out time was predetermined to 10 min. Therefore, the entire measurement time was 50 min.

### Venues

All investigated rooms are indoor event locations. To investigate the venue specific aerosol distribution patterns, venues with different room characteristics and ventilation systems (displacement ventilation, mixing ventilation and natural ventilation) were examined.

#### Displacement Ventilation Venue (DVV)

The investigated DVV is a rectangular theatre with 99 seats, an ascending spectator area and displacement ventilation. The air inlets are under the seats of row A to G and the air vents are on the upper left wall of the theatre (see Figure 3). Information about the room is presented in Table 1. The experimental measurements were performed without audience and heat sources were used to mimic thermal buoyancy.

**Figure 3:**
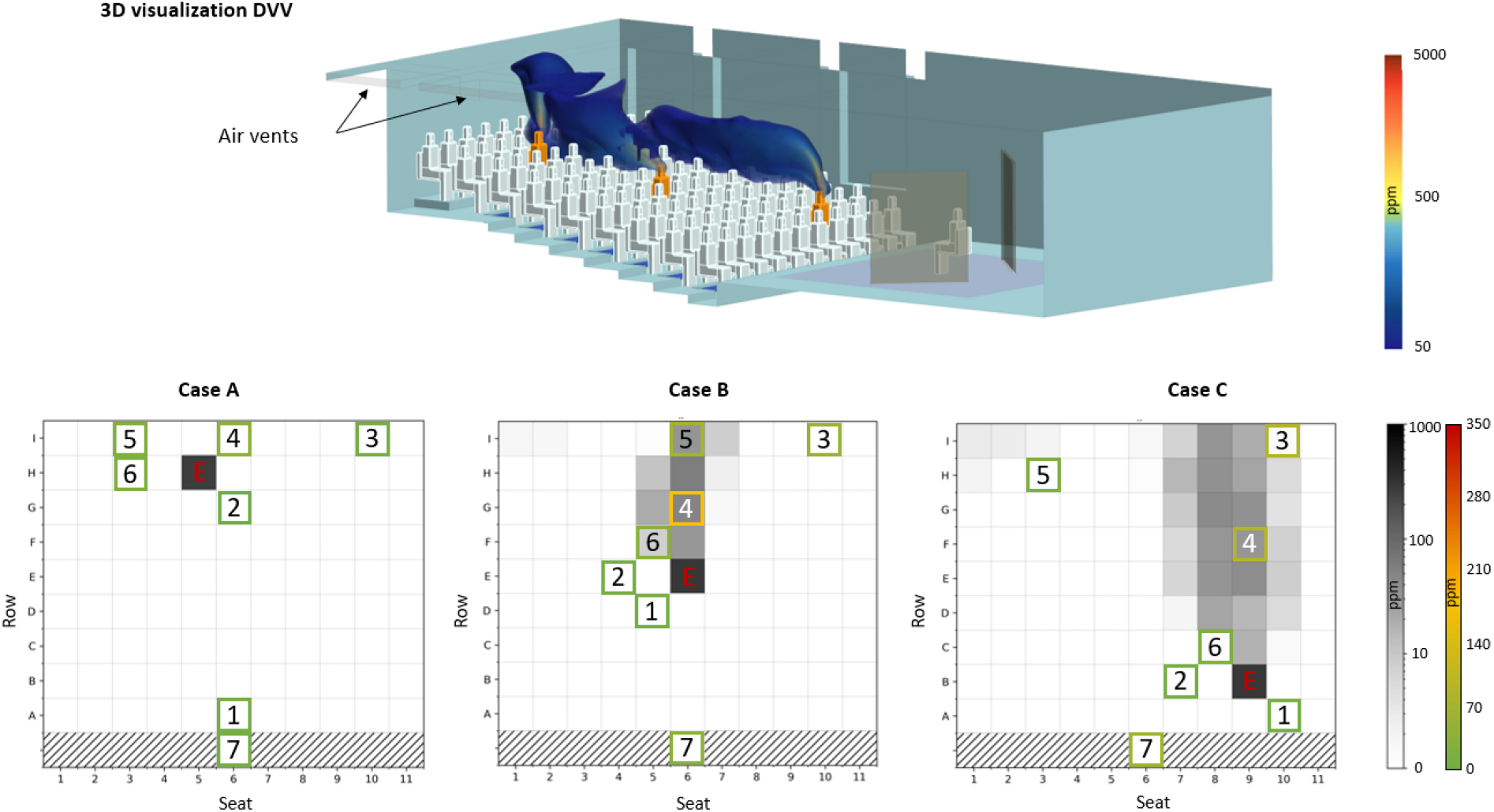
Experimental and simulated results of the three investigated emitter positions in DVV. In the 3D visualization the yellow dummies represent the positions of the aerosol generator measured with the experimental setup. Fresh air is inserted to the room under the seats and the polluted air is removed via air vents at the ceiling. The clouds over the yellow dummies are showing the aerosol plume dispersing in space calculated from numerical data. The three generator positions are: (Case A) aerosol generator in the back H5, (Case B) aerosol generator in the middle E6 and (Case C) aerosol generator in the front B9. E shows the position of the aerosol generator, number 1-7 the position of the absorbers. The colour of their frame indicates the measured aerosol amount. During the measurements heat sources were placed on the seats.

**Table 1:** Relevant parameter of the measurements in the DVV

| Quantity | Value | Quantity | Value |
| --- | --- | --- | --- |
| Ventilation type | displacement ventilation | Spectator area | ascending |
| Room volume | $530\text{ m}^3$ | Exposition time | 60 min |
| Seats | 99 | Audience | no |
| Ceiling height | 3.9 m | Date | 25.-27.10.2021 |
| Supply air volume flow | $4500\text{ m}^3/\text{h}$ | | |

#### Mixing Ventilation Venue (MVV)

The MVV has 103 seats, which are arranged in a three-stage arena shape. The audience is seated on the floor (see Figure 4). Further information is available in Table 2. The measurements were performed with heat sources to mimic an audience.

**Table 2:** Relevant parameter of the measurements in the MVV

| Quantity | Value | Quantity | Value |
| --- | --- | --- | --- |
| Ventilation type | mixing ventilation | Spectator area | two stage, arena shape |
| Room volume | 400 m<sup>3</sup> | Exposition time | 45 min |
| Seats | 103 | Audience | no |
| Ceiling height | 3.3 m | Date | 12.-13.01.2022 |
| Supply air volume flow | 6400 m<sup>3</sup>/h |  |  |

**Figure 4:**
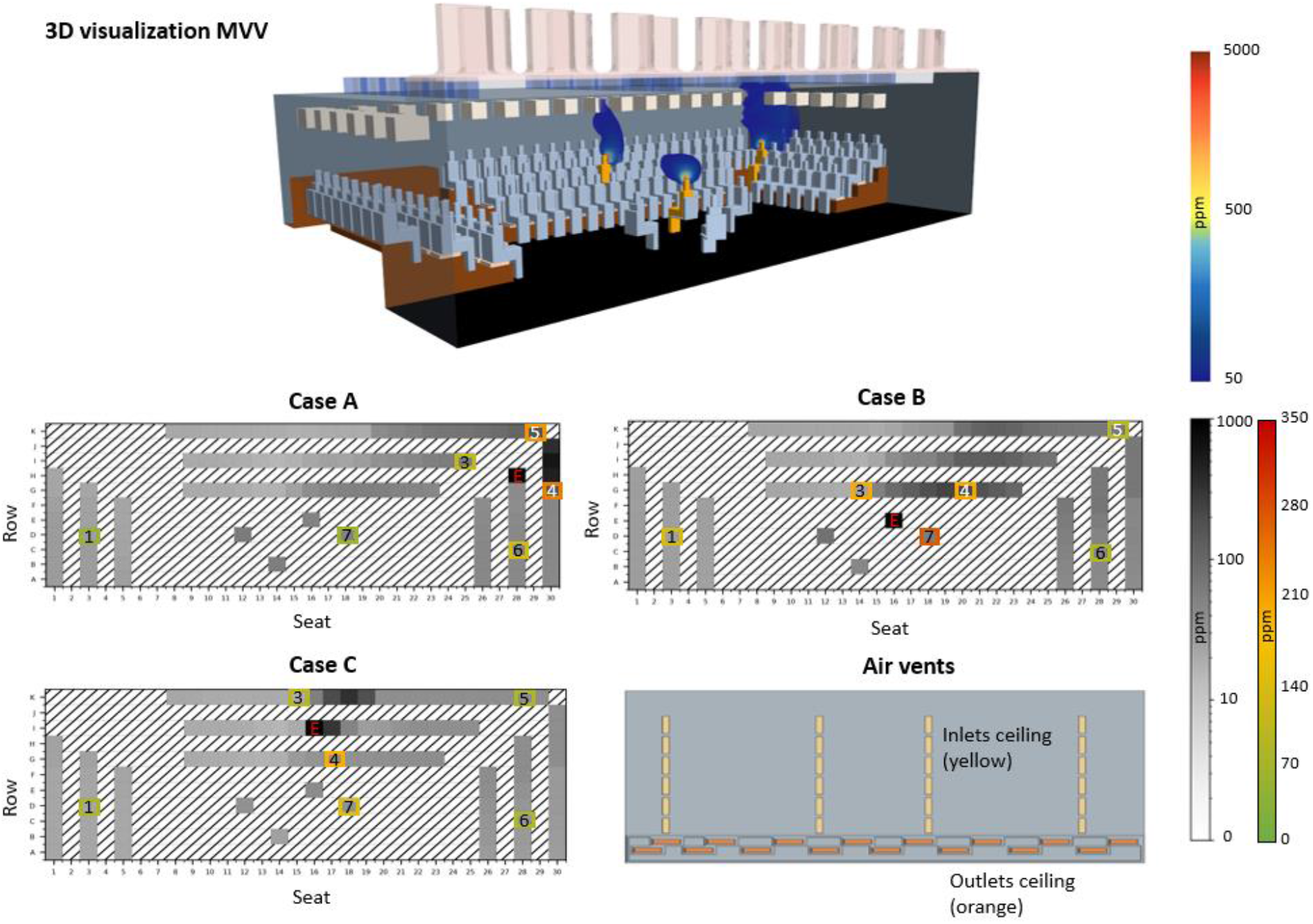
Experimental and simulated results of the three investigated emitter positions in MVV. In the 3D visualization the yellow dummies represent the positions of the aerosol generator measured with the experimental setup. A schematic representation of the air vents is shown (bottom right). The ventilation inlets are displayed in yellow and the ventilation outlets in orange. Both are located at the ceiling. The clouds over the yellow dummies are showing the simulated aerosol plume. The three generator positions are: (Case A) aerosol generator on the right H29, (Case B) aerosol generator on the stage E14 and (Case C) aerosol generator in the back I16. E shows the position of the aerosol generator, number 1-7 the position of the absorbers. The colour of their frame indicates the measured aerosol amount.

#### Natural Ventilation Venue (NVV)

In the venue with natural ventilation (NVV) air can only be exchanged through open doors and a smoke vent. Therefore, the air exchange is not quantifiable due to its strong dependence on thermal and external boundary conditions. The venue has 240 seats and an ascending spectator area (see Figure 5). Further information is available in Table 3. The measurement in this venue was conducted during a dress rehearsal with a real audience, so only one aerosol generator position could be investigated.

**Figure 5:**
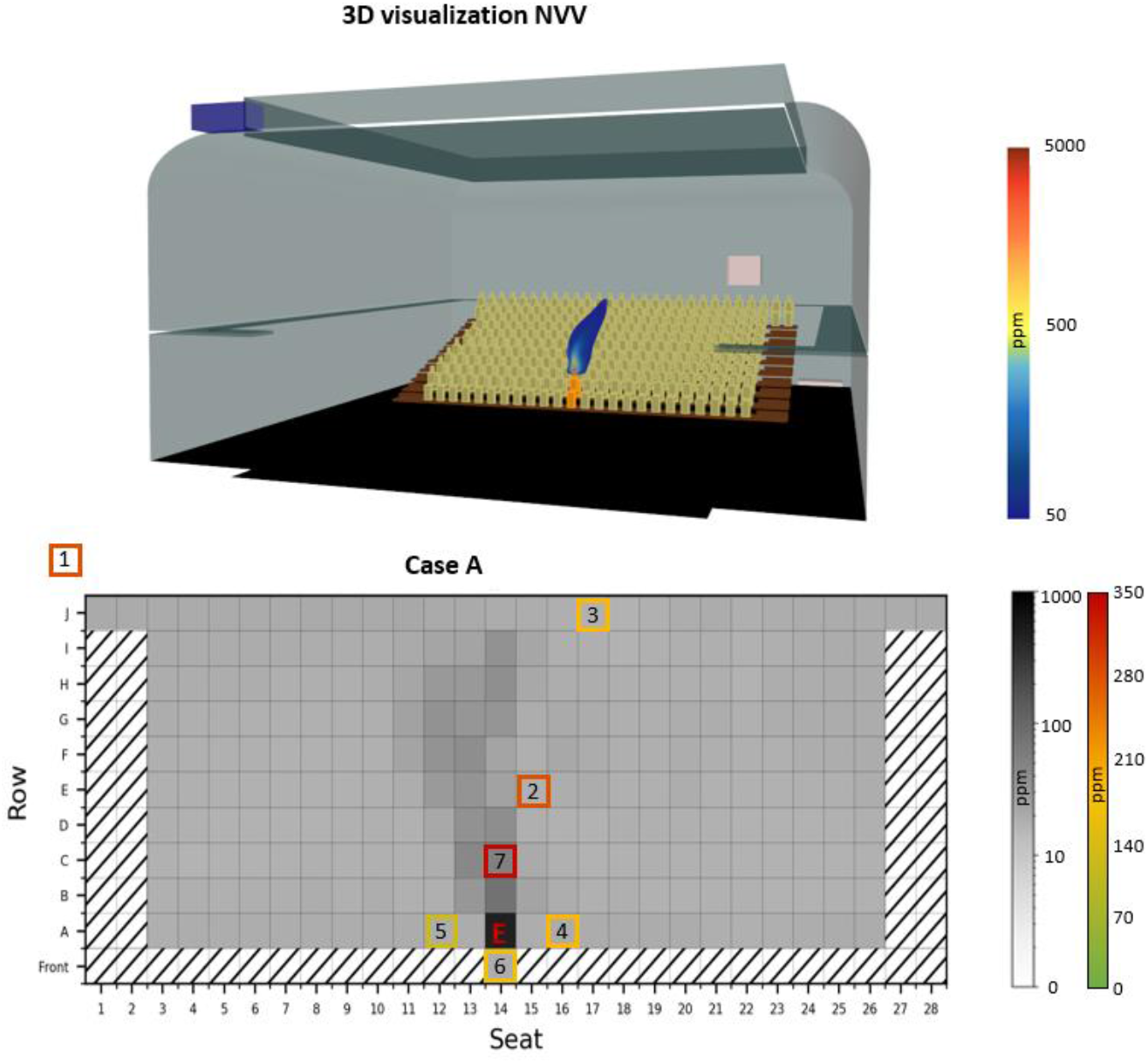
Experimental and simulated results of NVV. In the 3D visualization the yellow dummy represents the position of the aerosol generator measured with the experimental setup. The cloud over the yellow dummy is showing the aerosol plume dispersing in space calculated from numerical data. In Case A, the aerosol generator is positioned in the first row A14.

**Table 3:** Relevant parameter of the measurements in the NVV

| Quantity | Value | Quantity | Value |
| --- | --- | --- | --- |
| Ventilation type | natural ventilation | Spectator area | ascending |
| Room volume | 6000 m<sup>3</sup> | Exposition time | 30 min |
| Seats | 240 | Audience | yes |
| Ceiling height | 12.8 m | Date | 24.11.2021 |
| Supply air volume flow | unknown |  |  |

### Ethics Statement

The Ethics Committee of the Martin Luther University (Halle, Germany) approved the aerosol measurement experiments with the aerosol measurement system during regular events with spectators.

### CFD simulations

Non-isothermal, steady-state CFD simulations of the three venues were conducted on unstructured grids using Simcenter™ STAR-CCM+. The modelling of the venues closely followed constructions plans, walkthroughs, and additional information of the technical staff on site. Heat sources such as persons (assumed 80 W each), lighting and production equipment are remodeled to match the measurement configuration. Supply air and outside temperature are chosen based on the conditions at the day of the respective experiments. Mesh design relied on base sizes between 0.1 m and 0.3 m, with local refinements throughout the venue but especially close to heated or geometrically complex surfaces, and 4-6 prism layers were applied to support the near-wall modelling. In total, grid sizes ranged from 3.5 to 12 million finite volume cells. Air is treated as an incompressible, ideal gas where buoyancy effects are directly determined by the gravity model. Transport equations of flow and energy are solved with a segregated solver using the SIMPLE algorithm. Grey thermal, surface-to-surface radiation is used in combination with the segregated fluid temperature model. The Realizable K-ε-Model with a Two-Layer formulation (Wolfstein) is applied as a two-equation RANS-Model. Simplified mouth regions model the inlets of the tracer aerosol, where momentum-free passive scalars enter the fluid domain. For each emitting person (equivalent to aerosol generator) a separate passive scalar equation is solved. The volume-averaged concentrations of the respective tracer within the hemispherical volume around the absorber’s mouths (r= 0.23 m) are calculated and compared to the corresponding experimental value. Since the experiments relate the absorbed amount of NaCl to the emitted amount, a similar approach is chosen for the evaluation of the steady-state absorption of the tracer aerosol.

## RESULTS

The venue is displayed in a two-dimensional form as a matrix. To obtain aerosol distribution data for every position in the audience, CFD analyses were conducted. The numerical results are shown in shades of gray on a scale from 0 to 1000 ppm. The experimental results are obtained at seven selected seats by the measurement system. The results of the measurements are represented by a colour-coded outline of the seat on a color scale with values from 0 to 350 ppm.

### Reproducibility measurements of the system

Figure 2 shows the results of the reproducibility measurements. The mean values of the four absorbers are compared over time. 30 min after the start of the aerosol release a mean conductivity of μ = 4.92 μS/cm is detected with a standard variation of σ = 0.34. After 40 min, the mean conductivity is μ = 7.41 μS/cm with a standard variation of σ = 0.54. Thus, the signal-to-noise-ratio (SNR) after 30 min is:

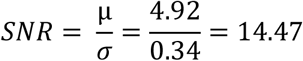

### Displacement ventilation venue

The influence of the positioning of the aerosol generator on the aerosol distribution was investigated in the DVV for the three cases A, B and C. In A the aerosol generator was placed in the back row (seat H5), in B it was placed in the middle row (seat E6) and in C in the second row of the spectator area (seat B9). Figure 3 and Table 4 show the results of this investigation. The position of the aerosol generator has a decisive influence on the distribution of aerosols in the venue. In the CFD data it can be seen that an aerosol plume forms behind the aerosol generator. The further forward in the audience the aerosol generator is placed, the more seats are affected by the plume and thus the higher the aerosol load in the venue. This trend can also be found in the experimental results.

**Table 4:** Experimental results of the investigations in the DVV.

| Absorber |  | 1 | 2 | 3 | 4 | 5 | 6 | 7 |
| --- | --- | --- | --- | --- | --- | --- | --- | --- |
| Aerosol amount [ppm] | A | 0 | 7 | 10 | 38 | 15 | 4 | 0 |
|  | B | 2 | 8 | 27 | 177 | 68 | 33 | 41 |
|  | C | 10 | 6 | 88 | 79 | 22 | 6 | 46 |

The experimentally derived amount of aerosol is highest in B and lowest in A (Figure 6). B and C show an increased measured aerosol amount at the positions directly behind the aerosol generator (absorbers 4, 5), while the positions next to (absorber 2) and in front (absorber 1) of the aerosol generator were characterised by a low aerosol absorption. In B and C, the absorber four seats behind the aerosol generator still absorbs around an order of magnitude more aerosol than the absorbers 1 and 2. The highest measured value can be found in the position two seats behind the absorber (B, absorber 4).

**Figure 6:**
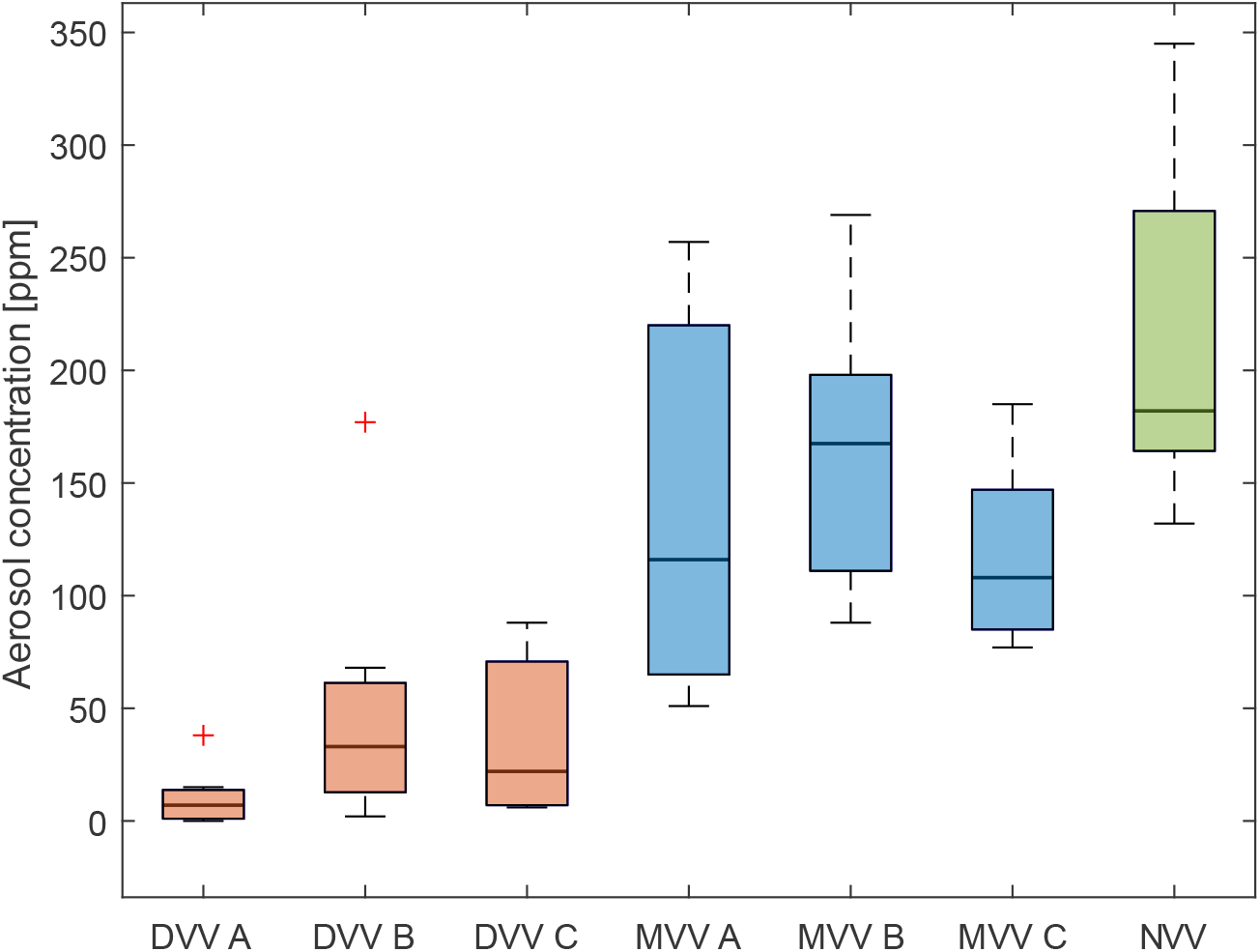
Comparison of the absorbed amount of aerosol at each measurement. The total aerosol load is significantly lower in DVV than in NVV. It should be noted that each boxplot consists of maximum 7 different measurement positions and therefore does not represent the total aerosol load in the room with sufficient accuracy. Red crosses represent outliers (outside 1.5 times the interquartile range).

### Mixing ventilation venue

The influence of the positioning of the aerosol generator on the aerosol distribution was investigated in the MVV for the three cases A, B and C. In A, the aerosol generator was placed on the right (seat H28), in B it was placed on the stage (seat E16) and in C in the back (seat I16). The results of this investigation are shown in Figure 4 and Table 5. In A, the highest aerosol amounts are measured by absorbers 4 and 5, the positions closest to the aerosol generator. Absorbers 3 and 6, which are also located near the aerosol generator, absorb a significant amount of aerosol. The same pattern can be observed for B and C. The three absorbers closest to the aerosol generator measure the highest amounts of aerosol. In both cases, the three absorbers placed further away from the aerosol generator together sampled about one third of the total aerosol measured. The total amount of measured aerosol by the six absorbers is the highest in B.

**Table 5:**
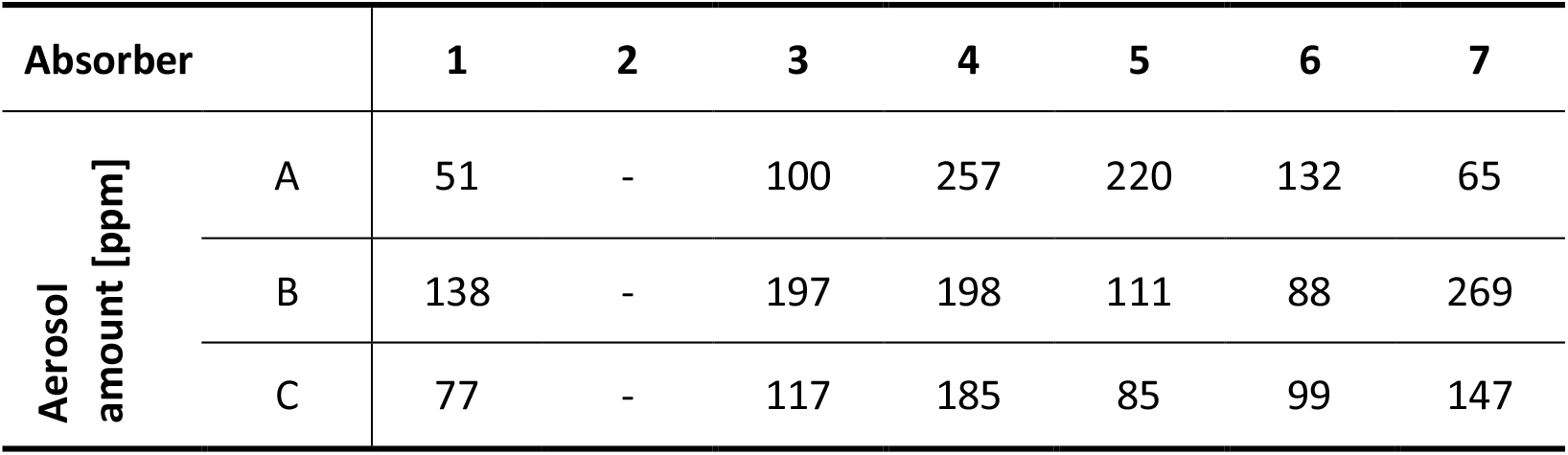
Experimental results of the investigations in the MVV. Malfunction of Absorber 2 during the measurements.

### Natural ventilation venue

For this venue only one position was examined, where the emitter is placed in the middle of the front row (seat A14). The results of this investigation are shown in Figure 5 and Table 6. The highest amount of aerosol was measured with the absorber 7 and 2 just behind the aerosol generator. Both the experimental and simulation data again show the formation of an aerosol plume, which is directed from the aerosol generator backwards over the audience. Even absorbers 1 and 3, which are located significantly further behind the aerosol generator, absorbed considerably more aerosol than the three absorbers in front (6) and next to the aerosol generator (4, 5). The latter absorbed only half as much aerosol as the most heavily exposed absorber 7 two seats behind the aerosol generator. Both the experimental and numerical results show that, in contrast to the DVV, the entire venue is highly loaded with aerosols. In the CFD there is no position with low amounts of aerosol.

**Table 6:** Experimental results of the investigations in the NVV.

| Absorber | 1 | 2 | 3 | 4 | 5 | 6 | 7 |
| --- | --- | --- | --- | --- | --- | --- | --- |
| Aerosol amount [ppm] | 270 | 271 | 182 | 177 | 132 | 160 | 345 |

**Table 7:** Summary of the characteristics of the three investigated venues based on experimental measurements and CFD analyses.

|  | **Displacement ventilation venue** | **Mixing ventilation venue** | **Natural ventilation venue** |
| --- | --- | --- | --- |
| **Aerosol distribution** | - Aerosol plume pulling backwards over the emitter | - No dominant room air flow, depending on boundary conditions<br>- Dispersion in the entire venue | - Aerosol plume pulling backwards over the emitter |
| **Positions with high aerosol exposure** | - Along the aerosol plume | - Forecast only possible to a limited extent | - Along the aerosol plume<br>- Prediction only possible to a limited extent |
| **Aerosol load of the venue** | - Low outside of the aerosol plume | - Medium, depending on the distance to the aerosol generator | - High in the whole venue and highest inside of the aerosol plume |

Notable is the high aerosol load measured by absorber 1. This absorber was placed outside of the spectator area slightly above the highest spectator rank at the level of theater lighting. Since the theatrical fog had accumulated as a layer predominantly at this height, it was considered an interesting position to measure the aerosol exposure.

### Comparison of the venues

In Figure 6 all measurements are shown in a boxplot. Even though a representative aerosol load cannot be determined from only seven measurement points in the room, a trend can be identified. The total aerosol load is significantly lower in DVV than in NVV. For a statistically meaningful conclusion, the CFD data should be considered here.

By comparing the dispersion characteristics of the aerosol in the different rooms, DVV and NVV show similar dispersion pattern of the aerosol plume. This aerosol plume forms behind the aerosol generator over the audience. Absorbers inside this plume measure multiple times higher aerosol loads compared to absorbers outside the plume. For the mixing ventilation a different dispersion pattern can be observed. A clear negative correlation between the measured aerosol and the distance to the emitter can be seen in Figure 7. All absorbers of the three MVV measurements are shown as a scatter plot. The aerosol amount decreases with increasing distance from the absorbers. This relation can be described with a power law (Figure 7 black dashed curve, f(x) = a*x^b^, a = 228.8, b= -0.418, R-square = 0.51).

**Figure 7:**
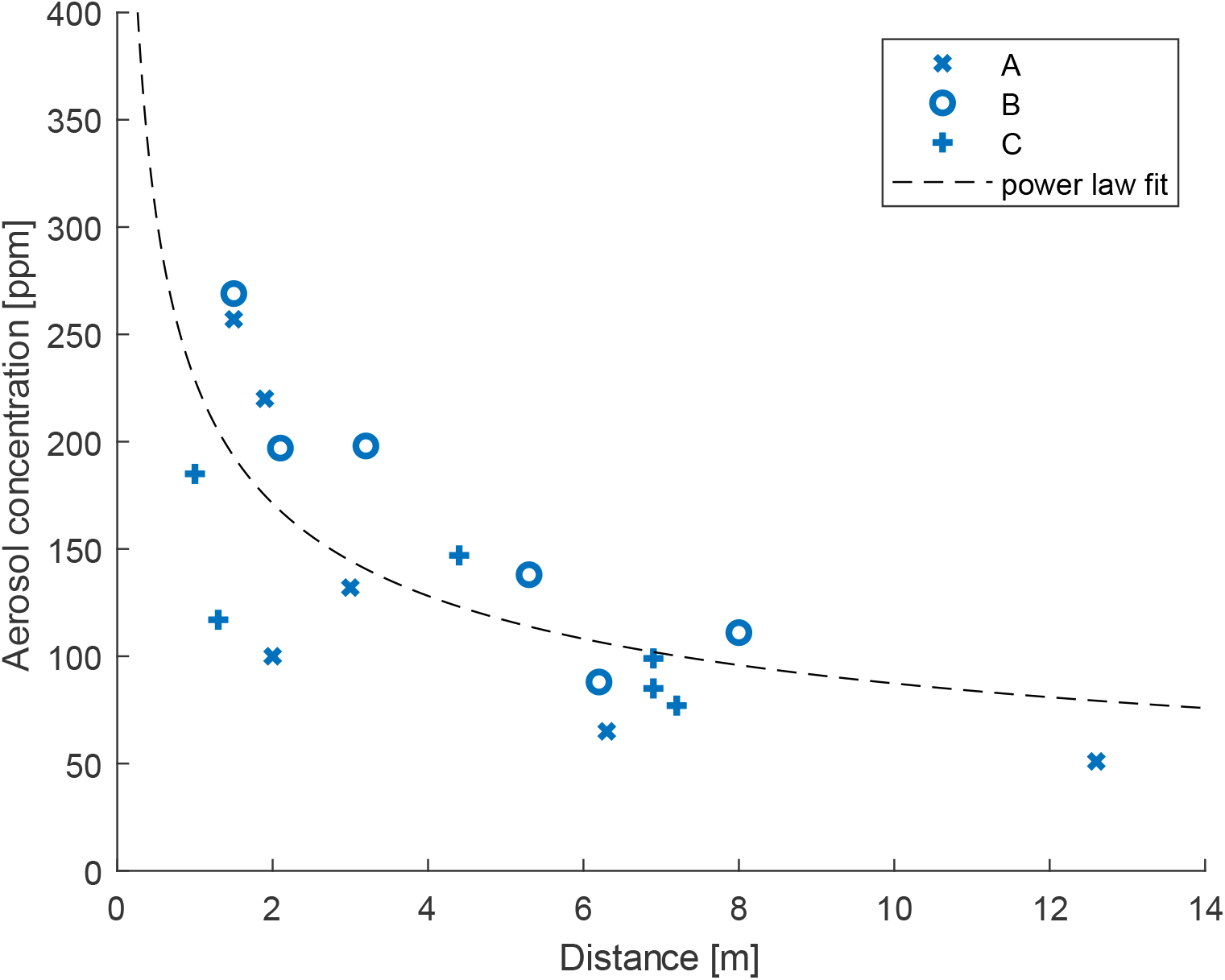
In the MVV, the aerosol amount decreases with increasing distance from the absorbers. This relation can be described with a power law (black curve, f(x) = a*x^b^, a = 228.8, b= -0.418, R-square = 0.51).

### Comparison of experimental measurements and CFD Simulations

Figure 8 Figure 7 shows the normalised amount of aerosol taken up by each absorber for both the experimental measurements and the CFD results. The consistency of data from experiments and simulations depends on the venue. For the DVV (orange) there are two groups of data points – the absorbers in the aerosol plume (upper right orange), and the absorbers outside the aerosol plume (lower left orange). For both groups, the data match quite well, although outside the aerosol plume slightly more aerosol was measured experimentally than numerically. The DVV values give an R-square value of 0.8633 when compared to the perfect match (black dashed diagonal). For the MVV, the data match less well than for the DVV. Across all three measurements, the match of the values deviates notably more than for DVV and NVV, with an R-square value of -0.1845. For the NVV, the experimental and simulation data match with an R-square value of 0.5155.

**Figure 8:**
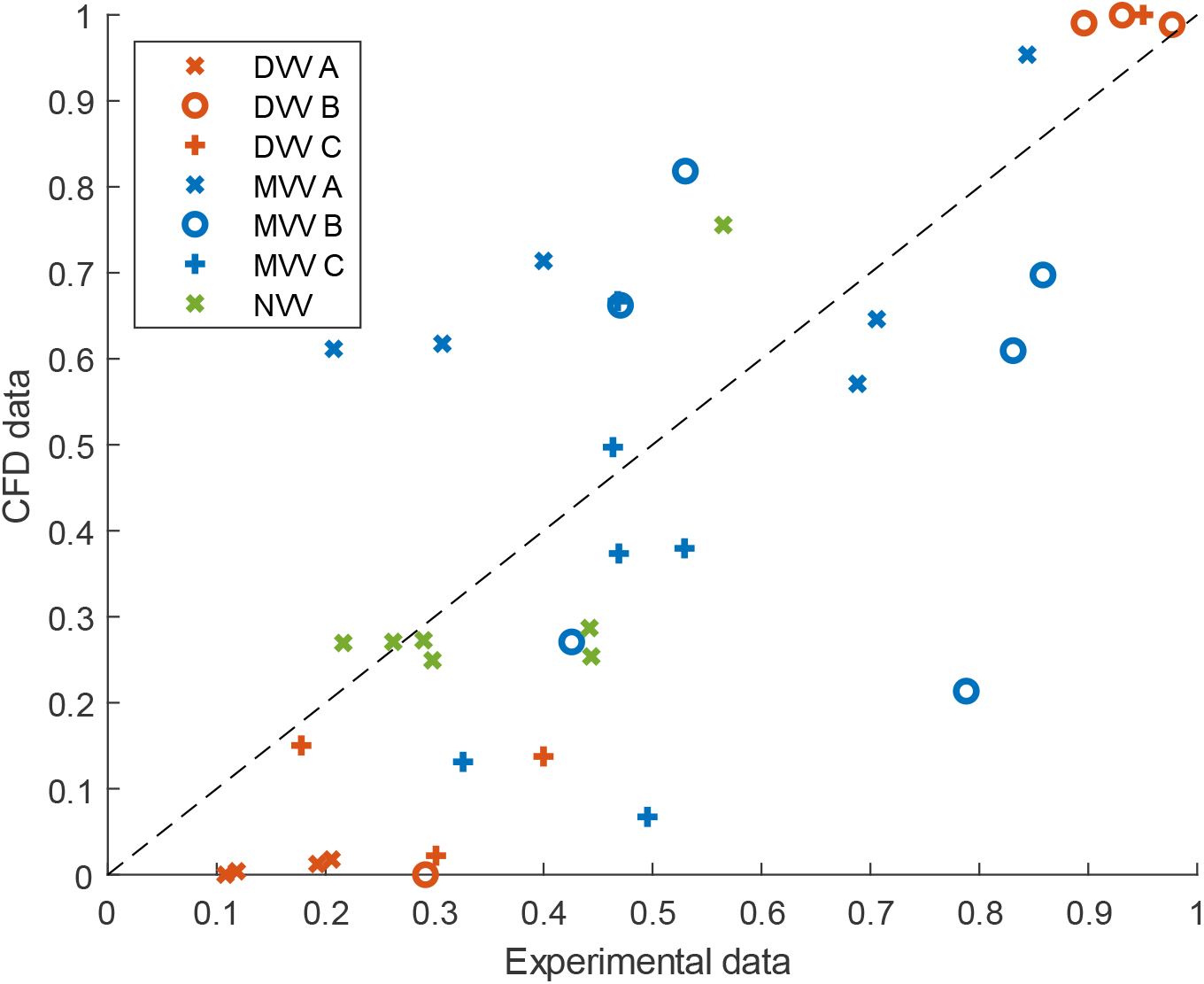
Comparison of normalized data points from CFD simulation and experimental measurements. Each colour represents a different venue. R-square values for the values compared to the perfect match (black dashed line): DVV R-square = 0.8633, MVV R-Square = -0.1845 and NVV R-Square = 0.5155.

## DISCUSSION

Case studies worldwide indicate that airborne aerosols are a major transmission route for SARS-CoV-2 virus indoors.^24,25^ So far, most COVID-19 risk evaluations for indoor environment are based on calculations applying spreadsheet models including relevant environmental and physiological parameters.^26,27^ Typical settings to evaluate the infection risk represent classrooms or office spaces with space volume less than 400 m^3^ and less than 50 persons present.^28^

The DVV with an ascending spectator area poses a good ventilation of the venue in all three scenarios, indicated by a low amount of absorbed aerosols. However, the observed aerosol plume was associated with highly exposed positions behind the aerosol generator, while the positions in front of and next to the aerosol generator remained almost unaffected, creating characteristic areas of low and high exposure. The position of the aerosol generator has a huge impact on the size of the aerosol plume and therefore on the aerosol exposure in the venue, as can be seen in Figure 3 between the rear position A and the front position C. The CFD data indicated that the further in the front of the ascending spectator area the aerosol generator is placed the higher the aerosol load in the entire venue. This can be explained by the positioning of the aerosol generator in proximity of the air vents (Figure 3-A), directly exhausting the emitted plume through the air vents. This observation is not supported by the experimental data, since in case C, a lower aerosol load is observed than in case B, despite a greater distance to the exhaust (Figure 6). However, this is possibly due to the unfortunate positioning of the absorbers. Only one absorber was positioned directly in the aerosol plume, so it is difficult to make a statement about the total aerosol load in the room.

It seems that the room air flow of the DVV can be reproduced well with CFD simulations. Qualitatively the distribution of aerosols in the simulation and the experiments match well. A closer look at the numbers shows that the high aerosol loads are overestimated by the simulation and the low ones are underestimated. However, numerical simulations seemed to adequately reproduce experimentally derived aerosol distribution patterns for DVV and are therefore a useful tool for aerosol distribution analysis when experimental measurements are not possible.

For the MVV, the differences of the absorbed amounts of aerosols between the aerosol generator positions only differ slightly. The total amount of aerosol measured by the absorbers was 77% higher compared to the DVV. In contrast to DVV and NVV, it was observed that the amount of aerosol decreases with increasing distance from the emitter, (Figure 7). This behaviour, which is shown in Figure 7, does not correspond to the common idea of an ideal mixing ventilation, in which the air is uniformly mixed throughout the room. However, predicting highly exposed positions is more difficult because the airflow in the mixing ventilation venue is less directional and seem to react sensitive to changing boundary conditions. This might also be the reason for the poor match of experimental and simulative data. It seems that the less predictable air flow in the MVV cannot be reproduced by the CFD simulation to match the experimental results. However, the higher aerosol load in the MVV compared to the DVV was shown in both experiments and CFD simulations.

The aerosol plume of the NVV behind the aerosol generator is wider and laterally less confined compared to DVV-C, which represents a similar emitter position. The measured amount of aerosol within the venue is much higher than the measured values at the other rooms, which is also confirmed by the CFD data. Similarly, current studies have shown that the risk of airborne transmission does not correlate with increasing distance in naturally ventilated venues, as the highest probability of infection were observed at greater distances.^29,30^ Even though there is no mechanical ventilation in the NVV, a very characteristic air flow is formed by the buoyancy above the spectators, which is also shown by CFD simulation, indicating good agreement between numerical and experimental data.

### Features and advantages of the measurement system

A major advantage of the measurement system is its wide range of applications. In contrast to other available measurement methods, it is not limited to clean rooms. The measurement system can perform in various sized and shaped indoor environments, with or without audience. If the measurements are conducted without an audience, heat sources are used, ensuring a higher reproducibility compared to measurements with a human audience. It needs to be emphasized that parameters such as room temperature, relative humidity and fluid turbulence influence the range and time aerosols are suspended in air.^24,28,31^ For these reasons, measurement results can vary with the slightest change in the boundary conditions. However, this also applies to dispersion of pathogenic aerosol in a real case. This paper focuses on fully occupied venues where the occupants are static most of the time. The measurement system can also be used in other ways, but the results are less reproducible if the air flow is influenced by a strong movement of people.

Since a NaCl solution is used as a tracer, the system is not harmful when used with an audience. A limitation might occur in environments where the air contains a certain concentration of conductive particles similar to the emitted tracer solution. Nevertheless, the background concentration of particles is measured anew for each measurement. Whether a room has been fully ventilated after a measurement or whether a background concentration is present can be deduced from a change in the increase in conductivity in the lead-in time, which is then subtracted from the total increase. To obtain the most meaningful measurement output, the positioning of the absorbers and aerosol generator at the venue is essential. Whether a tracer reaches a certain absorber in a certain time depends on the flow field. To get a good first impression of the global air flow for the application in rooms with complex airflow, it is helpful to perform a smoke dispersion analysis before the measurements.^32^ Subsequently, particularly critical areas can be analysed by the aerosol generator and the absorbers. The measuring system can also be used to investigate the influence of hygiene measures,^18^ to show the influence of the supply air volume flow and to show the effect of the occupancy of an venue.^19^

### Limitations

As the experimental measurement system only covers seven positions at the venue, it is currently not possible to representatively map the air flow of the room. It is difficult to select a representative configuration of the aerosol generator and absorber positions to cover the full range of sites with low and high aerosol amounts. Since the time for the measurements in the venues was limited for different reasons, our focus was to consider different aerosol generator positions to properly investigate the flow in the room. The reliability of the experimental data could be improved by using more absorbers and repeated measurements.

## CONCLUSION

In summary, the measuring system has shown that it is capable to examine venues of different sizes and characteristics making predictions about the aerosol distribution and aerosol load in the room. Due to its portability, it can be used on site. It can measure in real time. The experimental measurements allow for the examination of the effects of protective measurements like different ventilation strategies without underlying assumptions or restrictions like clean room environments.

The more the air flow in a venue is dominated by strong effects such as mechanical ventilation or thermal buoyancy, the better it can be reproduced with CFD simulations. For such venues, the experimental and simulative results showed good agreement. Here the experimental measurement system can offer a good possibility to validate CFD simulations. In venues with complicated undefined airflows, CFD simulations and many experimental measurement systems often cannot provide sufficiently reproducible data. For the fast and reliable evaluation of the dispersion of aerosols in such spaces, experimental systems such as our measurement system remain the primary option.

## Data Availability

The data that support the findings of this study are available from the corresponding author upon reasonable request.

## CONFLICT OF INTRESTS

The authors declare that there is no conflict of interest regarding the publication of this article.

## FUNDING STATEMENT

This work was supported by the Ministry of Science, Energy, Climate Protection and Environment of the Federal State of Saxony-Anhalt (grant number I 140), the Federal Government Commissioner for Culture and the Media (grant number 2521NSK115) and Berlin University Alliance (grant number SARS-CoV-2 Ausbreitung, Restart 2.0) as part of the RESTART 2.0 project.

## Notes

### Competing Interest Statement

The authors have declared no competing interest.

### Author Declarations

Ethics Committee of the Martin Luther University (Halle, Germany) gave ethical approval for this work.

